# Obesity, old age, and frailty are the true risk factors for COVID-19 mortality and not chronic disease or ethnicity

**DOI:** 10.1101/2020.08.12.20156257

**Authors:** Zinu Philipose, Nadia Smati, Chun Shing Jefferson Wong, Karen Aspey, Michael Mendall

## Abstract

Coronavirus-19 (COVID-19) mortality in hospitalized patients is strongly associated with old age, nursing home residence, male sex and obesity, with a more controversial association with ethnicity and chronic diseases, in particular diabetes mellitus. Further complicating the evaluation of the independent impacts of these risk factors is the failure to control for frailty in published studies thus far.

**Aim:** To determine the independent risk factors for mortality in patients confirmed to have COVID-19 needing hospital admission and to evaluate the independence of these risk factors after adjusting for body mass index (BMI) and frailty.

**Methods:** This observational study retrospectively reviewed hospital electronic medical records of 466 consecutive patients admitted to Croydon University Hospital confirmed positive by rapid PCR test from 11^th^ March to 9^th^ April 2020. Statistical analysis was performed by multiple unconditional and univariate logistic regression.

**Results:** After multivariate analysis, male sex [OR 1.44 (CI 0.92-2.40)], age per year [OR 1.07 (CI 1.05-1.09)], morbid obesity (BMI > 40 kg/m^2^ vs BMI 18.5-24.9 kg/m^2^) [OR 14.8 (CI 5.25-41.8)], and nursing home residence (OR 3.01 (CI 1.56-5.79) were independently associated with COVID-19 mortality with no statistically significant association found with chronic diseases or ethnicity. BMI was associated with mortality throughout its range from underweight to morbidly obese OR 1.07(1.03-1.11) per kg/m^2^ increase, p=0.0002 In the non-nursing home population, after adjusting for age and sex, the OR for type 2 diabetes mellitus (T2DM) as a risk factor was 1.64 (CI 1.03-2.61, p = 0.03) attenuated to 1.30 (CI 0.78-2.18)) after controlling for BMI.; the association of mortality with male sex was strengthened [OR 1.66 (CI 0.96-2.87)] and that for ethnic minority patients was weakened [South Asians [from OR 1.30 (CI 0.67-2.53)) to OR 1.21 (CI 0.60-2.46)]; Afro Caribbean [from OR 1.24 (CI 0.65-2.34) to OR 1.16 (CI 0.58-2.30)]. There was a borderline but potentially large protective effect (p= 0.09) of anticoagulation drug use prior to admission [OR 0.56 (CI 0.28-1.11)].

**Conclusion:** No significant effect of ethnicity and chronic diseases as independent risk factors on COVID-19 mortality was found whereas male sex, high BMI, old age, and frailty were found to be independent risk factors. BMI was related to mortality risk throughout its range from underweight to severely obese. It is likely that chronic diseases are epiphenomena of the effects of ageing and visceral adiposity on the immune system. Routine prophylactic treatment with anticoagulant drugs in the high risk COVID-19 population warrants further prompt investigation.

## Introduction

The Coronavirus-19 (COVID-19) pandemic of severe acute respiratory syndrome (SARS-CoV-2) has led to more than 580,000 deaths worldwide, as of July 2020.^1^ Increasing evidence reveals significantly higher hospital admission and death rates in males, the elderly, nursing home residents, ethnic minorities, the socially deprived, and patients with underlying diabetes, hypertension, cardiovascular disease, and obesity.^2^ However, the independence of each risk factor remains incompletely understood.

Two large scale studies in the United Kingdom (OpenSAFELY studies and the ISARIC CCP-UK) evaluated the characteristics and disease outcomes of hospitalized patients confirmed with COVID-19 infection by studying 17 million general practice patient records ^3^ and over 34,000 patient hospital records ^4^ respectively. Although both studies were large in scale, their methodology limited the characterization of the true effect of each risk factor, particularly the impact of obesity and frailty on mortality. The OpenSAFELY study employed information on body mass index (BMI) from general practice records up to 10 years before the COVID-19 infection^3^ and in the ISARIC CCP-UK study, the BMI was not objectively measured ^4^. In terms of frailty, although both studies included social deprivation indices related to locality, neither was able to explore the effects of individual frailty or the effect of nursing home residency. This could be a vital missing element since frailty has been suggested to be an independent risk factor for death from COVID-19 infection.^5^

Angiotensin-converting enzyme (ACE) inhibitors were initially proposed to have an influential effect on COVID-19 diagnosis and mortality but were found to have no statistically significant association later on.^6^ The proposed prothrombotic pathology of severe COVID-19 disease suggested that anticoagulant drugs may have a protective effect on disease outcome.^7^

Our aim, therefore, was to determine the true risk factors of COVID-19 mortality by studying a broad range of potential risk factors (both medical and social) in a consecutive case series of COVID-19 patient admissions to Croydon University Hospital, a district general hospital in the United Kingdom. In particular, we analysed the impact of ethnicity, contemporaneous BMI, frailty, nursing home residence, pre-existing chronic diseases, co-existing medication history, and smoking history on inpatient mortality.

## Methods

### Study design, patients and data collection

This single centre retrospective observational cohort study included all patients admitted to Croydon University Hospital from 11^th^ March to 9^th^ April 2020 and had confirmed COVID-19 infection by Polymerase Chain Reaction (PCR) nasopharyngeal antigen swab test. Data collection ceased on 1^st^June 2020 by which time, all patients had an outcome as either deceased or discharged. These patients had respiratory symptoms and signs of COVID-19 infection on admission as criteria for testing during this period. The patients’ hospital information on Cerner electronic medical records were reviewed by a team of physicians who then collected their epidemiological data, past medical history, medication history, clinical and laboratory data, and outcomes from their admission to either their discharge from hospital or death. An anonymous identification code was assigned to each patient record to protect patient confidentiality. All electronic data were stored in and analysed on hospital servers. According to the guidelines of the UK Health Research Authority, as the data were collected as part of routine clinical care and were evaluated retrospectively, the study was considered a review of clinical practice and ethical approval was not required. This study was conducted in accordance with the Declaration of Helsinki.

### Outcomes

The primary outcome was mortality.

### Definition of Covariates

COVID-19 symptom severity was categorized based on patients’ oxygen requirements and complications throughout admission. Patients were classified in the mild symptom category if their room-air oxygen saturation levels were maintained at or above 93%; moderate symptom category if they had a respiratory rate above or equal to 30 breaths per minute and/or room-air oxygen saturation below 93%; and severe symptom category if they developed respiratory failure and/or multi-organ failure.

BMI was ascertained from the height and weight recorded on the COVID-19 infection admission but if unavailable, the most recent recording from the last six months prior to the infection. Ethnic group categories were established from admission clerking and patient registration details. These were grouped as White/Caucasian, South Asian, Black/Afro Caribbean, East Asian, Middle Eastern/North African (MENA), and Undetermined. Patient’s residence was categorised as unassisted living residence (such as their own home, family’s home, or rented accommodation) or an assisted living residence (such as care homes, nursing homes, and sheltered accommodation). Dependence on activities of daily living was whether or not a patient required carers’ help for daily activities. Occupation was categorised as manual, non-manual work, or retired/none. Smoking history was categorised as current, former, or none. History of alcohol excess was categorised as yes if consumption was above 14 units per week.

In terms of patients’ medical background of pre-existing chronic diseases, we examined their current and previous respiratory, cardiac, liver, vascular, cerebrovascular and immunocompromised conditions. Respiratory history included any respiratory comorbidities such as obstructive or restrictive pulmonary disease and/or history of tuberculosis. Cardiac history included a history of ischemic heart disease and/or arrhythmias. Immunocompromised states included active cancer, Human Immunodeficiency Virus and/or on immunosuppressive medications (including high dose corticosteroids, biological agents, systemic chemotherapy medication). Arterial and venous vascular diseases included a history of hypertension, cerebrovascular events, venous clots including pulmonary embolism and deep venous thrombosis. We also examined if patients had type 2 diabetes mellitus (T2DM) or type 1 diabetes mellitus (T1DM).

Patients’ drug history was based on their regular medication that was recorded on admission. The following medications were recorded: Statin, Metformin, antihypertensives, antiplatelets and anticoagulants. Statin doses were equalized into Atorvastatin dose equivalents. The use of any antihypertensive drug was recorded but we specifically isolated the use of ACE-1 inhibitor drugs and angiotensin receptor blocker drugs for further analysis.

Laboratory markers (serum alanine transaminase (ALT), vitamin D, albumin, C-reactive protein (CRP), creatinine kinase (CK), Haemoglobin (Hb), Troponin-T, Haemoglobin A_1_C (HbA_1_C)) were analysed. Patients’ ALT results were recorded both pre-admission and on admission. Patients’ pre-admission vitamin D levels were recorded. HbA_1_C was only recorded if the measurement was taken less than six months prior to admission. All of the other markers were taken on admission. Liver ultrasound records were examined up to five years prior to admission looking for reports of cirrhosis or fatty liver disease.

### Statistical Analysis

We used Statview SE for statistical analysis. Univariate statistical analysis comparisons for death or discharged alive were performed using Chi-square test for two level categorical variables of the following: sex, type of residence, dependency for activities of daily living; history of T1DM, T2DM, cardiac disease, stroke, thromboembolism, respiratory disease; the use of statins, metformin, anti-hypertensives, antiplatelet agents, anticoagulants; and ultrasound evidence of fatty liver or cirrhosis.

Univariate logistic regression was used for age groups (<45, 45-59, 60-69, 70-79, 80-89, 90+years), body mass indexes (0-18.5, 18.6-25, 26-30, 31-40, <40 kg/m^2^, and unknown), ethnic groups (Caucasian, South Asian, Afro-Caribbean, East Asian, Middle Eastern/North African, and undetermined), disease severity during admission (mild, moderate, severe), smoking history (current, former, none), excess alcohol history (yes/no), and occupation (manual, non-manual, retired/none). In a separate analysis BMI was also modelled s a continuous variable amongst the subjects for whom values were available.

The Mann-Whitney U test was used to compare continuous explanatory variables that were not normally distributed: age, BMI, HbA_1_C, biomarkers on admission (CRP, ALT, troponin-T, and CK) and pre-admission (Hb and Vitamin D).

Multivariate analysis was performed with unconditional logistic regression and the following variables were retained on the final model: age was taken as a continuous variable; sex, ethnic group, BMI were taken as categorical variables; the following were taken as binary variables (yes/no): nursing home residence, dependence for activities for daily living, history of T2DM, cardiac disease, respiratory disease, stroke, hypertension and the use of anticoagulants. Tests of significance were derived from likelihood ratio tests.

## Results

We analysed 466 consecutive subjects (59% male and 41% female). 267 patients were discharged and 199 patients died.

The pre-admission characteristics of patients who were discharged or died before and after adjustment for age and sex of subjects are described in Table 1. On univariate analysis, age and the morbidly obese BMI category (>40 kg/m^2^) were powerfully associated with mortality risk, and this remained after adjustment for age and sex [OR 13.2 (CI 5.10-34.9), p<0.0001], with lowest risk being in the underweight. When BMI was modelled as a continuous variable, after adjustment for age and sex the OR per kg/m2 increase was 1.06(1.03-1.09) p=0.0004. More modest associations with cardiac and hypertension history and antiplatelet agents disappeared after adjustment for age and sex, but treatment with anticoagulants emerged as a borderline protective factor. Of the indicators of frailty, dependence on activities of daily living and nursing home residence were powerfully associated with risk of mortality with 29% of all deaths in hospital related to COVID-19 being nursing home residents, but the effect of dependence for activities of daily living disappeared after adjustment. There was only a very weak non-significant association with ethnic group apart from a higher mortality in those grouped as “undetermined”.

**Table 1:** Pre-admission patient characteristics and their univariate associations with death

| Characteristics | Discharged (n= 267) | Died (n= 199) | p-value | Odds ratio for mortality (95% CI) adjusted for age and sex only (n=466) | p-value |
| --- | --- | --- | --- | --- | --- |
| **Continuous age** (years)<br>(median, IQR, Range) | 67 (26, 6-97) | 77 (17,43-98) | NS | 1.05 (1.03-1.06) | <0.0001 |
| **Categorical age** (years) |  |  | <0.0001 |  | <0.0001 |
| <45 | 34 (12.7%) | 2 (1.0%) |  | 1 |  |
| 45-59 | 68 (25.5%) | 22 (11.1%) |  | 5.61 (1.24-25.3) |  |
| 60-69 | 46 (17.2%) | 43 (21.6%) |  | 16.1 (3.64-71.2) |  |
| 70-79 | 54 (20.2%) | 41 (20.6%) |  | 13.4 (3.04-59.3) |  |
| 80-89 | 51 (19.1%) | 69 (34.6%) |  | 24.1 (5.50-105) |  |
| 90+ | 14 (5.3%) | 22 (11.1%) |  | 28.1 (5.79-136) |  |
| **Sex: Male** | 158 (59.2%) | 119 (59.8%) | NS | 1.28 (0.86-1.90) | NS |
| **Ethnic Group** |  |  | NS |  | NS |
| White/Caucasian | 119 (44.6%) | 93 (46.7%) |  | 1 |  |
| South Asian | 52 (19.5%) | 29 (14.7%) |  | 1.10 (0.62-1.94) |  |
| Afro-Caribbean | 59 (22.1%) | 35 (17.8%) |  | 1.17 (0.68-2.02) |  |
| East Asian | 4 (1.5%) | 4 (2.0%) |  | 1.94 (0.45-8.42) |  |
| MENA | 2 (0.7%) | 2 (0.5%) |  | 1.22 (0.11-14.0) |  |
| Undetermined | 31 (11.6%) | 36 (18.3%) |  | 2.20 (1.20-4.02) |  |
| **BMI** (kg/m<sup>2</sup>) |  |  | <0.0001 |  | <0.0001 |
| 0-18.5 | 15 (5.6%) | 6 (3.0%) |  | 0.84 (0.27-2.56) |  |
| 18.6- 25 | 76 (28.5%) | 48 (24.1%) |  | 1 |  |
| 26-30 | 49 (18.4%) | 49 (24.6%) |  | 2.07 (1.17-3.70) |  |
| 31- 40 | 70 (26.2%) | 27 (13.6%) |  | 1.20 (0.64-2.25) |  |
| >40 | 10 (3.7%) | 18 (9.1%) |  | 13.2 (5.10-34.9) |  |
| Unknown | 47 (17.6%) | 51 (25.6%) |  | 3.20 (1.70-6.03) |  |
| **Dependence for ADLs** | 93 (34.8%) | 99 (49.7%) | <0.002 | 1.10(0.71-1.71) | NS |
| **Residence** |  |  | 0.002 |  | 0.008 |
| Unassisted home | 230 (86.1%) | 141(70.8%) |  | 1.95(1.18-3.21) |  |
| Assisted home | 35 (13.1%) | 58 (29.1%) |  |  |  |
| **Occupation** |  |  | 0.0005 |  | NS |
| Non manual | 40 (15.0%) | 15 (7.5%) |  | 0.43 (0.11-1.70) |  |
| Manual | 17 (6.4%) | 3 (1.5%) |  | Ref |  |
| Retired/None | 184 (68.9%) | 174(87.4%) |  | 1.20(0.59-2.47) |  |
| **Excess Alcohol History** | 31 (11.6%) | 10 (5.3%) | NS | 0.82(0.40-1.70) | NS |
| **Smoking History** |  |  | NS |  | NS |
| Former | 204(76.5%) | 137 (68.8%) |  | Ref |  |
| None | 44 (16.6%) | 33 (16.6%) |  | 0.90 (0.53-1.51) |  |
| Current | 19 (7.1%) | 9 (4.7%) |  | 0.93 (0.39-2.21) |  |
| **Medical background** |  |  |  |  |  |
| Type 1 diabetes mellitus | 3 (1.1%) | 3 (1.5%) | NS | 2.12 (0.36-12.3) | NS |
| Type 2 diabetes mellitus | 83 (31.0%) | 76 (38.2%) | 0.09 | 1.38 (0.92-2.07) | 0.12 |
| Hypertension | 123 (46%) | 111 (55.7%) | 0.047 | 1.17 (0.79-1.73) | NS |
| Cardiac history | 78 (29.2%) | 79 (39.7%) | 0.02 | 1.38 (0.92-2.07) | 0.12 |
| Stroke History | 29 (10.8%) | 26 (13.0%) | NS | 0.70 (0.38-1.29) | NS |
| Thromboembolism | 18 (6.7%) | 12 (6.0%) | NS | 1.03(0.65-1.59) | NS |
| Respiratory History | 77 (28.8%) | 54 (27.1%) | NS | 0.97 (0.63-1.49) | NS |
| Immunosuppression | 32 (12%) | 34 (17%) | 0.1 | 1.27 (0.74-2.20) | NS |
| **Drug history** |  |  |  |  |  |
| Statin | 85 (31.8%) | 79 (39.6%) | NS | 1.03 (0.71-1.45) | NS |
| Metformin | 55 (20.6%) | 45 (22.6%) | NS | 1.39 (0.84-2.16) | NS |
| ARBs | 25 (9.4%) | 20 (10.0%) | NS | 0.89 (0.47-1.70) | NS |
| ACEi | 48 (18%) | 35 (18%) | NS | 0.97 (0.59-1.58) | NS |
| Other antihypertensives | 85 (32%) | 77 (39%) | NS | 1.12 (0.75-1.68) | NS |
| Antiplatelets | 48 (18%) | 51 (26%) | 0.052 | 1.03 (0.64-1.67) | NS |
| Anticoagulants | 39 (15%) | 29 (15%) | NS | 0.59 (0.33-1.03) | 0.06 |
| **Biochemical markers**<br>(median, IQR, range, count) |  |  |  |  |  |
| HbA<sub>1c</sub> (mmol/mol) | 45(15, 21-153, 103) | 50.5 (23.5, 0-108, 68) | 0.02 | 1.02(0.99-1.03) | 0.06 |
| ALT ((IU/L)) pre-admission | 24(17, 7-121, 212) | 18 (12.3, 0-113, 141) | <0.0001 | 0.99(0.97-1.003) | NS |
| Vitamin D (nmol/L) | 30(33, 0-100, 61) | 35 (39, 13-113, 37) | NS | 1.00(0.99-1.02) | NS |
| **Liver Ultrasound** |  |  | NS |  | 0.09 |
| Normal/None | 238 (89.1%) | 188 (94.5%) |  | Ref |  |
| Fatty liver | 26 (9.7%) | 11 (5.5%) |  | 0.57(0.27-1.25) |  |
| Cirrhosis | 3 (1.1%) | 0 (0.0%) |  | ----- |  |

BMI = Body mass index, ADL= Activities of daily living, ARB= Angiotensin receptor blockers, ACEi= Angiotensin converting enzyme inhibitors, HbA<sub>1c</sub>= Haemoglobin A<sub>1c</sub>, ALT= Alanine aminotransferase

The distribution of elderly (age 61 years and above) within each ethnic group was as follows: 68.2% (144/211) White/Caucasian, 40.7% (33/81) South Asian patients, 41.5% (39/94) Black/Afro-Caribbean patients, and 42.3% (33/78) MENA/East-Asian/Other/Undetermined patients. The distribution of obesity (BMI 30 kg/m^2^and above) within each ethnic group was as follows: 27.8% (50/180) White/Caucasian, 33.3% (20/60) South Asian (rising to 39% if a BMI cut off for obesity of 27.5 kg/m^2^ is used (8–11), 45.3% (34/75) Black/Afro-Caribbean patients, and 39.6% (21/53) of MENA/East-Asian/Other/ Undetermined patients.

The relation of laboratory values on admission to risk of in hospital mortality is listed in Table 2. CRP was higher and albumin lower. Troponin was only available on 142 subjects but showed a strong association with mortality. Unsurprisingly, mortality was much higher in those subjects with severe disease on admission compared to those with moderate or mild disease.

**Table 2:** Patient characteristics during admission and their univariate associations with death

| Characteristics | Discharged (n = 267) | Died (n = 199) | p-value |
| --- | --- | --- | --- |
| **Disease Severity** |  |  | <0.0001 |
| Mild | 57 (21.3%) | 1 (0.5%) |  |
| Moderate | 170 (63.7%) | 10 (5.0%) |  |
| Severe | 40 (15.0%) | 188 (94.5%) |  |
| **Biochemical markers on admission**<br>(Median, IQR range, count) |  |  |  |
| CRP | 68 (106.5, 2.1-450, 197) | 125 (150, 2-606, 197) | <0.0001 |
| Albumin | 32 (6, 12-46, 258) | 29 (6, 14-40) | <0.0001 |
| Creatinine Kinase | 128 (225, 0-4659, 41) | 304 (970, 12-27225, 35) | 0.01 |
| Hb | 131 (27, 79-180, 267) | 124 (27, 71-186) | 0.001 |
| Troponin-T | 14 (18.3, 0-358, 81) | 39 (37, 3-1091, 61) | <0.0001 |
| ALT | 29.5 (31.5, 5-337, 260) | 26 (30, 7-795, 183) | NS |

CRP= C reactive protein, Hb= Haemoglobin, ALT= alanine aminotransferase

The multivariate analysis of pre-admission characteristics associated with COVID-19 mortality is shown in Table 3. There were strong and independent associations with age, nursing home residence and BMI. The effect of BMI was mainly in subjects with BMIs greater than 40 [OR 14.8(CI 5.25-41.8), p<0.0001], with the lowest risk being in underweight subjects. There was evidence of a continuous positive association with BMI in the 375 subjects with data OR 1.07(1.03-1.11) per kg/m^2^ increase, p=0.0002. Weaker associations were observed with male sex, and a possible protective effect of anti-coagulant treatment. There were modest and non-significant associations with ethnic minority groups.

**Table 3:** Multivariate comparisons of determinants of mortality

| Variable | Odds ratio(95%CI)<br>mutually adjusted<br>n=466 | p-value<br>(for heterogeneity) | Odds ratio mutually adjusted excluding nursing home residents<br>n=358 | p-value<br>(for heterogeneity) |
| --- | --- | --- | --- | --- |
| **Age** (per year) | 1.07 (1.05-1.09) | <0.0001 | 1.07(1.05-1.10) | <0.0001 |
| **Sex: Male** | 1.44 (0.92-2.40) | 0.13 | 1.66(0.96-2.87) | 0.07 |
| **Ethnic group** |  | NS |  | NS |
| Caucasian (reference) | 1 |  | 1 |  |
| South Asian | 1.30 (0.67-2.53) |  | 1.21(0.60-2.46) |  |
| Afro Caribbean | 1.24 (0.65-2.34) |  | 1.16(0.58-2.30) |  |
| East Asian | 2.94 (0.59-14.7) |  | 2.69(0.51-14.1) |  |
| MENA | 0.88 (0.04-22.3) |  | 0.70(0.02-23.4) |  |
| Undetermined | 1.98 (0.99-3.97) |  | 1.51(0.65-3.52) |  |
| **BMI(kg/m<sup>2</sup>)** |  | <0.0001 |  | <0.0001 |
| <18.5 | 0.62 (0.18-2.10) |  | 0.39(0.04-3.71) |  |
| 18.5-25 reference | 1 |  | 1 |  |
| 26-30 | 1.22 (1.18-4.08) |  | 2.09(1.00-4.36) |  |
| 31-40 | 1.38 (0.70-2.69) |  | 1.50(0.70-3.21) |  |
| 40+ | 14.8 (5.25-41.8) |  | 17.7(5.63-55.4) |  |
| missing | 3.35 (1.67-6.71) |  | 3.35(1.54-7.30) |  |
| **Nursing home resident** | 3.01 (1.56-5.79) | 0.0008 | ----- | ----- |
| **Dependent of ADL** | 0.94 (0.54-1.63) | NS | 0.93(0.52-1.67) | NS |
| **T2DM** | 1.28 (0.79-2.07) | NS | 1.35(0.78-2.31) | NS |
| **Cardiac history** | 1.30 (0.76-2.21) | NS | 1.61(0.87-3.00) | 0.13 |
| **Respiratory history** | 0.74 (0.45-1.23) | NS | 0.61(0.34-1.09) | 0.09 |
| **Stroke history** | 0.65 (0.32-1.32) | NS | 0.57(0.23-1.40) | NS |
| **Hypertension** | 1.00 (0.63-1.58) | NS | 1.02(0.60-1.72) | NS |
| **Anti-coagulants** | 0.56 (0.28-1.11) | 0.09 | 0.48((0.21-1.15) | 0.09 |

BMI = Body mass index, ADL= activities of daily living, T2DM= Type 2 Diabetes Mellitus,

The nursing home population had a median age of 82 (range 51-96) versus 66 [range 6-98 (p<0.0001)] for the remaining population. 29% (28/96) in the nursing home population versus 35% (129/364) in the non-nursing home population were diabetic. Only 15% (14/82) were obese or morbidly obese versus 32% [116/289 (p<0.0001)] in the others. Excluding the nursing home population had varying effects on associations with mortality: the associations with BMI and diabetes were strengthened while the association with ethnicity was weakened. In the non-nursing home population after age and sex adjustment the OR for T2DM mortality was 1.64 [CI 1.03-2.61 (p=0.03)]. This was not attenuated after controlling for ethnic groups, OR 1.59 [CI 0.99-2.58 (p=0.06)], but was markedly attenuated after further controlling for BMI, OR 1.30 (CI 0.78-2.18). In the nursing home population, there was no association between T2DM and mortality [OR 1.22 (CI 0.48-3.15)] which was not significantly altered after controlling for ethnic group [OR1.10 (CI 0.39-3.14)] or BMI [OR 1.19 (CI 0.38-3.70)].

## Discussion

In our retrospective observational study of 466 consecutive patients hospitalized with confirmed COVID-19 disease, for a defined period from 11, March 2020 to 9, April 2020, the overall death rate 24.5%. Strong associations were observed with BMI. Underweight subjects had the lowest risk and the morbidly obese had the highest risk. Age and nursing home residence were also strongly associated. The relationship with male sex was strengthened and of borderline significance after excluding nursing home residents. There was a protective effect of borderline statistical significance of a potentially important magnitude of anticoagulant therapy. Ethnicity was only weakly associated with risk which was further attenuated after excluding nursing home residents. None of the chronic diseases were significantly associated with mortality. The association with T2DM was present only after excluding nursing home residents but attenuated markedly after adjusting for BMI. Higher CRP, CK and Troponin-T values on admission showed strong associations with mortality.

The strengths of our study are the detailed information on current BMI and the inclusion of measures for frailty. A number of other observational studies involving patients admitted with COVID-19 have confirmed the marked association of mortality with BMI ^8–11^ although our findings are at the upper end of the range and none have commented on the protective value of being underweight. It is of value to compare the findings from current study with two other larger studies from the United Kingdom (UK). The OpenSAFELY study used general practice medical records and was able to study the risk associated with ethnic groups versus the general population.^3^ It however could not control for frailty and relied on any BMI recorded within the past 10 years. This may explain why only modest but statistically significant associations were observed with BMI. Similar associations to the present study of only a very modest magnitude with mortality were observed with ethnic groups, and T2DM explained some of this. However, they were unable to control as adequately for BMI and frailty which explained much of the association of T2DM with mortality in the current study. The ISARIC CCP-UK study also from the UK was of similar design to ours, including subjects from several centres and hence larger. It however again could not adequately control for BMI, relying on observation as to whether simply the subjects were obese or not and again included no measures of frailty. Their patient population had a different ethnic composition (83% were White/Caucasian versus our study was 51% White/Caucasian) but similar very modest associations were seen with ethnic groups, partly explained by T2DM.^4^

A study from New York of risk factors for admission with COVID19 and the development of critical illness identified obesity as a risk factor but its effect on mortality was not specifically addressed.^8^ A smaller study from the same city again identified obesity as a risk factor for mortality in those with a BMI >35 kg/m^2^ with BMI 25-34.9 kg/m^2^ as the reference group.^10^ In our study when redefining the reference range in this manner, we get a similar magnitude of effect for BMI > 35 kg/m^2^. These studies did not explore the effect of being underweight or of frailty.

We demonstrated only modest associations of ethnic groups with mortality which fell short of statistical significance yet were of a similar magnitude to those observed in the OpenSAFELY and ISARIC studies. No associations with ethnic groups were found in the studies from New York.^8,10^ Moreover, in many countries where some of the minority ethnic population groups originate from, the overall COVID-19 mortality rate per 1 million population has not shown to be higher than in the UK.^1^ This suggests there is no obvious genetic predisposition between ethnic groups for COVID-19 pathology susceptibility, and that social factors may go some way in explaining the Black, Asian, and minority ethnic (BAME) findings in the United Kingdom.^12^

In the present study, the exclusion of nursing home residents weakened the association between ethnic groups and mortality. Similarly, the association between T2DM and mortality was also attenuated after controlling for BMI, which was previously postulated as part of the mechanism for the association of ethnic groups with mortality.^3,4^ We suspect therefore that residual confounding may be part of the explanation of the apparent association of ethnic groups with mortality of admitted patients.

In the local Croydon population, the White British group comprises 55.1%, South Asians 16.4%, and Afro-Caribbeans 20.2%.^13^ On the surface this is similar to the proportion of the different ethnic groups admitted to hospital in our study. However, patients from ethnic minorities were younger on average, and made up a disproportionately large number of admitted patients aged <60 years, suggesting that they are more likely to develop severe COVID-19. This is likely a result of being more at risk of being infected with COVID-19 due to behavioural and socio-economic factors rather than the development of more severe COVID once infected, as per the findings of Petrilli et al. in New York.^8^

It seems likely that if objective measures of visceral adiposity were utilized, we would be able to elucidate much more clearly the apparent relationships between the presence of T2DM, ethnicity and COVID-19 outcomes. South Asians in particular are prone to developing increased visceral adiposity at lower BMIs, and according to national guidelines, are considered obese with a BMI of 27.5kg/m^2^.^14–17^ We have previously demonstrated that the inflammatory response to a defined injury increases markedly with waist: hip ratio as well as age, with a threefold higher peak C-reactive protein response in the top tertile versus the lowest tertile.^18^ This is likely to result from the pro-inflammatory state associated with visceral obesity which plays an important role in the pathogenesis of severe COVID-19.

The World Health Organization has stated nearly half of the COVID-19 related deaths in Europe by June 2020 have been from the nursing home population.^19^ Nursing home residence is likely to be a good marker of frailty according to most definitions.^20^ There is a paucity of data on the impact of being a resident in a nursing home on other potential mediators of mortality in COVID-19 and we were able to study this in more detail. The nursing home population were less likely to have conventional risk factors including obesity and T2DM than age matched hospitalized patients. We therefore hypothesize that frailty per se is likely to be the driving factor for mortality in COVID-19 nursing home resident patients beyond obesity, diabetes and their associated comorbidities. On the other hand, patients who were dependent for their activities of daily living were not found to be at a higher risk for mortality if they were not residing in nursing homes. We also cannot preclude that levels of care were different for the elderly patients from nursing homes versus from non-nursing homes. Additionally, frailty could be acting through other mechanisms and may represent enhanced inflamm-aging.^21^ Nonetheless, it is important to account for nursing home residence and frailty when studying the risk factors for COVID-19 mortality.

The role of male sex is a consistent predictor of mortality in all COVID-19 studies.^22^ We demonstrated male sex as an independent risk factor for COVID-19 mortality in our analysis.

Research findings have proposed theories based on the X chromosome’s roles in immune response activation ^23^, the Angiotensin-converting enzyme-2 ^24^, and testosterone modulation of Transmembrane Serine Protease 2.^25^ Further work both examining biological and social gendered differences is required to understand this difference in pathogenicity between male and females. Additionally, we cannot exclude that men are more prone to acquiring COVID-19 as male sex was over-represented amongst admissions compared to the local population. It also may be that if men are infected, they are more likely to have severe symptoms and present to hospital. This can be resolved by sero-epidemiology studies on the community.

The possible marked protective effect of anticoagulants, which our study was underpowered to detect conclusively, is in concordance with what we are learning about the pathogenesis of severe COVID-19 and the prominent role that thrombosis plays.^26^ There were no other trends found for use of antihypertensives or antiplatelet agents. There was also no significant effect of other chronic disease after adjustment for age and sex. It is therefore likely that associations with other chronic comorbidities are confounded by age and visceral adiposity.

One challenge of our study was missing patient data. We have done our best to include these cases in our analyses. The size of the study precluded us from exploring in more detail the effect of medication history on prognosis and detecting some effects of pre-existing chronic disease. Despite this, our study has identified the most important and clinically significant pre-morbid conditions influencing mortality. Our study captured data from the first 466 consecutive patients admitted to Croydon University Hospital and therefore the earlier time period of the pandemic in the UK, so there have been significant changes in understanding and management of the condition since our data analysis. The fact that we captured data during the peak of patient admission burden for COVID-19 meant there were significant pressures on the Intensive Care Unit, affecting selection criteria to more level 3 care on a daily basis. This may have influenced some patient outcomes. Furthermore, during this time period, COVID-19 testing was in short supply and therefore, not every patient who was admitted was tested, particularly those with mild or no symptoms.

## Conclusion

Obesity, or possibly more specifically abdominal adiposity, as well as male sex, ageing, and frailty are likely to be the key factors in determining the outcome of COVID-19 infection. Further studies are required to prove that other postulated risk factors such as ethnicity and chronic disease, including diabetes, are independent risk factors, but we could find no evidence. It is likely that these are epiphenomena of the effects of visceral adiposity and aging on the immune system. Prophylactic anticoagulation in high risk subjects where COVID-19 is widely prevalent could be life-saving and warrants further study.

## Data Availability

All our data is anonymized and available from a password protected secure location in keeping with the hospital trust guidelines and can be made available upon formal request.

## Acknowledgements

We thank the Research and Development team and the Microbiology team for their support in this project.

We also would like to express our gratitude to Dr Alok Mehta, Dr Georgios Karanasios, Dr Eugene Yap, Dr Joseph Hogan, Dr Homira Ayubi, and Dr Amy Woods for their contributions to this study

